# Time-series modeling of epidemics in complex populations: detecting changes in incidence volatility over time

**DOI:** 10.1101/2025.02.19.25322557

**Authors:** Rachael Aber, Yanming Di, Benjamin D. Dalziel

## Abstract

Trends in infectious disease incidence provide important information about epidemic dynamics and prospects for control. Higher-frequency variation around incidence trends can shed light on the processes driving epidemics in complex populations, as transmission heterogeneity, shifting landscapes of susceptibility, and fluctuations in reporting can impact the volatility of observed case counts. However, measures of temporal volatility in incidence, and how volatility changes over time, are often overlooked in population-level analyses of incidence data, which typically focus on moving averages. Here we present a statistical framework to quantify temporal changes in incidence dispersion and detect rapid shifts in the dispersion parameter, which may signal new epidemic phases. We apply the method to COVID-19 incidence data in 144 United States (US) counties from the January 1st, 2020 to March 23rd, 2023. Theory predicts that dispersion should be inversely proportional to incidence, however our method reveals pronounced temporal trends in dispersion that are not explained by incidence alone, but which are replicated across counties. In particular, dispersion increased around the major surge in cases in 2022, and highly overdispersed patterns became more frequent later in the time series. These findings suggest that heterogeneity in transmission, susceptibility, and reporting could play important roles in driving large surges and extending epidemic duration. The dispersion of incidence time series can contain structured information which enhances predictive understanding of the underlying drivers of transmission, with potential applications as leading indicators for public health response.

**Author summary:** Understanding patterns in infectious disease incidence is crucial for understanding epidemic dynamics and for developing effective public health policy. Traditional metrics used to quantify incidence patterns often overlook variability as an important characteristic of incidence time series. Quantifying variability around incidence trends can elucidate important underlying processes, including transmission heterogeneity. We developed a statistical framework to quantify temporal changes in case count dispersion within a single time series and applied the method to COVID-19 case count data. We found that conspicuous shifts in dispersion occurred across counties concurrently, and that these shifts were not explained by incidence alone. Dispersion increased around peaks in incidence such as the major surge in cases in 2022, and dispersion also increased as the pandemic progressed. These increases potentially indicate transmission heterogeneity, changes in the susceptibility landscape, or that there were changes in reporting. Shifts in dispersion can also indicate shifts in epidemic phase, so our method provides a way for public health officials to anticipate and manage changes in epidemic regime and the drivers of transmission.

## Introduction

Time series of infectious disease incidence appear, to varying degrees, “noisy”, showing higher frequency fluctuations (e.g., day-to-day or week-to-week fluctuations) around trends at the broader temporal ranges typical for epidemic curves (e.g., months or years). Short-term fluctuations in incidence time series are caused in part by variable reporting, but may also reflect the population-level impacts of transmission heterogeneity and changes in the landscape of susceptibility [1, 2, 3, 4, 5, 6, 7, 8]. Metrics of variability in incidence time series may therefore carry information regarding underlying drivers of transmission, and offer a relatively unexplored avenue for understanding epidemic dynamics.

Contact tracing data has revealed temporal changes in the variability of individual reproductive numbers, quantified by shifts in the dispersion parameter of the offspring distribution in branching process models [7, 8]. Similar evidence has been recovered through statistical reconstruction of transmission networks, indicating temporal trends in the level of dispersion at different phases of an epidemic [3]. However, the scaling from individual-level transmission heterogeneity to population-level epidemic dynamics is not fully understood. In addition, traditional contact tracing is very resource intensive, and although new approaches using digital technologies may improve its speed and scalability [9], it would be helpful to have complementary population-level analyses that can estimate heterogeneity using incidence data, which is more widely available. The importance of considering population-level variability and its relationship to individual-level variability is further highlighted by the finding that a combination of individual-based and population-based strategies were required for SARS-CoV-2 control during the early phases of the pandemic in China [6]. An important challenge therefore is to develop methods that can detect changes in population-level variability in incidence time series, and to interpret these changes in terms of underlying transmission processes.

Emerging statistical techniques are leveraging variability in epidemic time series to enhance understanding of disease dynamics at the population level. For example, a recently-developed method uses population-level incidence data to estimate the dispersion parameter of the offspring distribution, which quantifies heterogeneity in secondary cases generated by an infected individual [5]. It is also possible to estimate the dispersion parameter of the offspring distribution from the distribution of the final size of a series of localized outbreaks [10]. Clustering of cases has also been estimated directly from incidence data [11]. Another important application links variability in inci-dence to epidemic phases; for example, changes in the mean and interannual coefficient of variation of measles incidence have been used to identify a country’s position on the path to elimination, providing insights into vaccination strategies and epidemiological dynamics [12]. Analysis of the shape of epidemic curves for influenza in cities may identify contexts where incidence is focused more intensely (proportionally more infections in a smaller span of time) with implications for the sensitivity of cities to climate forcing and for surge capacity in the health system [4, 13].

What drives incidence dispersion and how does it relate to the underlying branching process of transmission, and to observations of cases? Under a wide range of configurations for a branching process model of contagion spread, the number of infected individuals *I*_*t*_ at time *t* will have a negative binomial distribution [14, 15], *I*_*t*_ ~ *NB* (*µ*_*t*_, *θ*_*t*_), where *µ*_*t*_ is the expectation for *I*_*t*_ and *θ*_*t*_ is the dispersion parameter. The variance is related to the mean and dispersion parameters by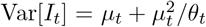, so smaller values of the dispersion parameter *θ*_*t*_ correspond to increasing amounts of dispersion, which increase the amounts by which the variance in realized number infected *I*_*t*_ exceeds the expected value, *µ*_*t*_. Conversely, the distribution of *I*_*t*_ tends to a Poisson distribution (where the variance equals mean) as *θ*_*t*_ becomes large. The negative binomial distribution may also accurately model a time series if there is a changing process mean within a time step: for example, if the mean of a Poisson distribution itself follows a gamma distribution, the resulting distribution is negative binomial. Negative binomial regression (in contrast to Poisson regression) can account for unobserved heterogeneity, time dependence in the rate of a process and contagion within a time step that all lead to overdispersion [16].

An interpretation of the dispersion parameter for a time series model of counts is that events are 1 + *θ*^−1^ times as “crowded” in time relative to a Poisson process with the same mean [17] (see derivation in S1 Text). For example, *θ* = 1 corresponds to a situation where the average number of infections in the same time step as a randomly selected case will exceed the Poisson expectation by a factor of two. In a simple example relevant to surge capacity in healthcare systems, *θ* = 1 implies that a random infectious individual visiting the emergency department at a hospital would find it on average to be twice as crowded with other infectious individuals (infected by the same pathogen) as expected for a Poisson process with the same incidence rate.

In a sufficiently large host population, and when the infectious pathogen can be assumed to spread in nonoverlapping generations, the number of infections each generation is often modeled as

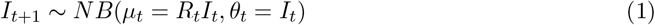

where time-varying reproductive number *R*_*t*_ gives the expected number of secondary infections acquired from an infected individual at time *t*, and the generation time is set to 1 without loss of generality [14, 18]. Setting *θ*_*t*_ = *I*_*t*_ arises from the assumption that individuals who acquire the infection at time *t* form independent lineages with identically distributed local rate parameters. In applications, this model for *theta* becomes *θ*_*t*_ = *C*_*t*_*/ρ*_*t*_ where *C*_*t*_ represents reported cases and *ρ* the reporting rate, which relates reported cases to the true number of infections as *C*_*t*_ = *ρ*_*t*_*I*_*t*_. However, this requires that susceptible depletion in one lineage does not affect another, that transmission rates are equal across lineages, and that reporting rates do not vary across lineages.

In practice, these assumption will not often hold, and our aim in this paper is to develop, test and apply an alternative approach which produces data-driven estimates of *θ*_*t*_, including identifying timepoints when *θ* is changing rapidly, which may help to reveal the impacts of heterogeneity in transmission, susceptibility, and reporting.

## Methods

By definition incidence volatility is fast relative to broadscale epidemic dynamics.

Consequently, in order to estimate incidence volatility we first modeled incidence at broad spatiotemporal scales using natural splines [19]. To allow for diverse shapes in the broadscale epidemic dynamics, spline modeling was conducted within a moving window such that for each half of the window

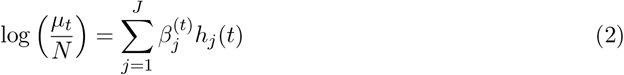

where *N* represents population size, *h*_*j*_(*t*) are basis functions, the degrees of freedom is equal to the number of knots *k* for the natural spline, *J* = *k* + *d* + 1, where *d* is the degree of the polynomial, and 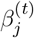 are fitted parameters. The window has half-width Δ, centered at *t*, i.e., extending from *t* − Δ to *t* + Δ. The degrees of freedom (number of knots) to be used for the splines, and the width of the moving window will depend on the application. Explanation of the specific choices we used for our application to COVID-19 cases in US counties is described below.

Modeling the underlying epidemic dynamics based on log-transformed incidence allows us to address the statistical effects of population size on the relationship between the mean and variance in count data, which would otherwise confound our analysis. Specifically, since population size influences the mean and variance of case count data, it impacts dispersion in different-sized populations that are otherwise identical. Accordingly, population size appears as an offset in our model of broad-scale incidence changes. That is,

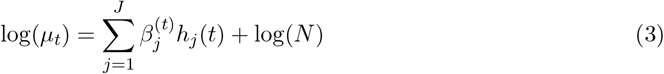

The form of the probability mass function (PMF) for infections at a time step is:

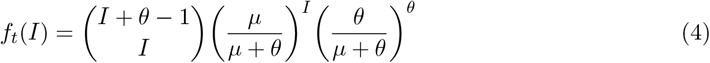

where *µ* is estimated via the linear predictor outlined above.

We estimate *θ*_*t*_ from observed incidence data using an iteratively reweighted least-squares (IRLS) procedure for mean estimation, combined with the optimize function in R, which uses a combination of golden section search and successive parabolic interpolation, to compute *θ*_*t*_. Specifically, within each time window, the spline model with an offset term was used to estimate a series of *µ*_*s*_ values for *s* = *t* − Δ to *s* = *t* + Δ via IRLS, as implemented in the NBPSeq R package [20]. A single value of *θ*_*t*_ was then calculated for the entire time window by maximizing the likelihood function, which is based on the negative binomial probability mass function defined above.

In addition to fitting the model at each time step, we developed a likelihood-ratio test (LRT) to test the hypothesis that *θ* has changed at each time step. This test involves fitting and comparing two models: a null model (no *θ* change) and a two-part model (with a *θ* change). For the null model, a single *θ* value was fitted for the entire time window. For the *θ*-change model, separate *θ* values were fitted for the left (from *t* − Δ to *t*) and right (from *t* to *t* + Δ) halves of the time window.

Very large *θ* values correspond to processes that are operationally identical to a Poisson process. Accordingly, the test does not produce a *p*-value if any of the three *θ* estimates exceed a user-specified threshold. In the application below, we set this threshold at 10^3^, meaning that *θ* estimates with temporal crowding within 0.1% of that expected for a Poisson process were considered effectively Poisson.

Similarly, values of *θ* very close to 0 focus all of the mass of the PMF on 0, representing a scenario where the probability of observing any infections approaches zero. As with the Poisson-like tolerance described in the previous paragraph, our algorithm does not produce a *p*-value if any of the three *θ* estimates are below a user-specified threshold. This threshold will depend on the presence of contiguous sections of the time series being analyzed during which no cases are observed. In the application below, we set this threshold to 10^−3^, because *θ* values below this level correspond to 0 frequencies that greatly exceed those in the data.

With both upper and lower *θ* thresholds–—corresponding to Poisson-like and zero tolerances, respectively—–maximum likelihood estimates (MLEs) of *θ* beyond these thresholds exhibited unbounded behavior. When *θ* exceeded the upper threshold, corresponding to processes operationally identical to a Poisson process, the MLE tended to grow arbitrarily large, with the likelihood function reaching its maximum at the upper boundary of the calculated domain. Conversely, when *θ* fell below the lower threshold, representing extreme overdispersion with probability mass concentrated near zero, the MLE approached zero, and the likelihood function peaked at the lower boundary of the domain. This behavior reflects the inability of the model to reliably estimate *θ* when it lies outside the specified thresholds (Fig 1).

**Fig 1.**
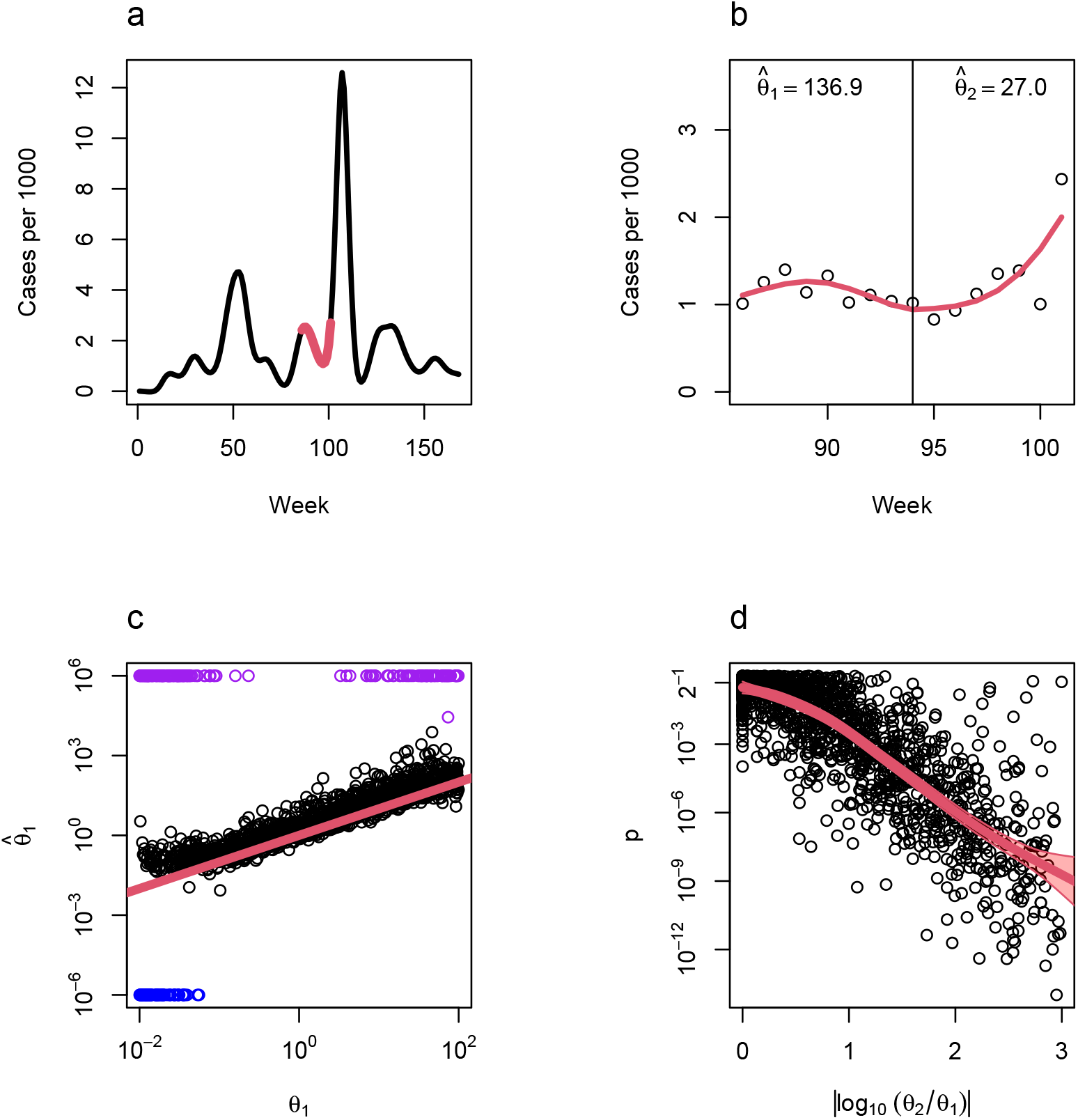
Detecting dispersion changes in case count time series. a: Weekly incidence of COVID-19 in the United States, with time measured in weeks since January 4, 2020, showing an example of a randomly-selected 16 week period used as an incidence trend when in simulation-based validation of the LRT test (red). b: Cases in one county (Douglas County, Nebraska) over the sample time period with estimated incidence trend (red) and estimated dispersion values on either side of the midpoint. c: Estimated *θ* versus true *θ* in simulation studies combining a randomly-selected section of the national incidence curve with a random population size and set of dispersion values. Estimated values outside of tolerance plotted in purple (close to Poisson) and blue (close to collapsing to zero). d: Statistical power of the LRT test.

### Application to simulated data

We evaluated the robustness of our framework to a range of population sizes, magnitudes of dispersion changes, and shapes of underlying incidence trends by generating 2,000 simulated epidemic curves with known parameters. Epidemic trends were modeled as smoothed incidence series derived from 16-week sections randomly selected from US COVID-19 data (described below), scaled to reflect different population sizes ranging from 10^3^ to 10^7^. For each simulated trajectory, dispersion parameters (*θ*_1_ and *θ*_2_) were assigned to the two halves of the selected 16-week window, and case counts were simulated using a negative binomial distribution, where the mean (*µ*) was based on the smoothed incidence trend scaled by the population size. The values of *θ*_1_ and *θ*_2_ were drawn from a uniform distribution spanning 10^−2^ to 10^2^, with 10% of simulations set to have no change in dispersion (*θ*_1_ = *θ*_2_). Extremely large differences in dispersion (absolute log-ratio *>* 3) were capped by setting *θ*_2_ = *θ*_1_.

### Application to empirical data

We applied our framework to COVID-19 case data for the United States at the administrative level of counties, compiled by The New York Times, based on reports from state and local health agencies between Jan 4, 2020, and March 18, 2023 [21], and using county population sizes estimated for 2021 from the United States Census Bureau [22]. Cumulative cases for the largest three counties in each state were converted to weekly counts by keeping the last observation from each week and differencing to compute new cases. Occasionally, reported cumulative case counts were not monotonically increasing due to corrections posted by local agencies as they resolved incoming data. As a result, approximately 0.24% of estimated new cases across all counties in the dataset were negative and these were set to zero. For each county, we analyzed overlapping 16-week windows, shifting one week (i.e., one timestep) at a time. Within each window, the framework estimated the dispersion parameter (*θ*) using a natural spline with three degrees of freedom for each half of the window to model the broad-scale trend in incidence. Outputs included estimated dispersion parameters (*θ*_1_, *θ*_2_, for the left and right halves of each window, and *θ* for the entire window), likelihood ratio test statistics, *p*-values for changes in dispersion at the midpoint of the window, and flags for boundary conditions such as failure to reject Poisson-like dispersion or collapse to extreme overdispersion.

## Results and Discussion

Simulations indicate that the LRT framework accurately detects changes in dispersion, with p-values converging to 0.5 as the effect size approaches 0, reflecting the uniform distribution of p-values under the null hypothesis, and decreasing toward zero as the effect size increases (Fig 1). The framework is also robust to the range of population sizes present in the empirical data—county populations ranged from approximately 48 thousand to 9.9 million, and we tested the framework on simulated populations between 10 thousand and 10 million. Across this range, the method produced accurate estimates for *θ* within 10^−2^ ≤ *θ* ≤ 10^2^, encompassing all operationally relevant values for COVID-19 incidence data and many other infectious diseases. Lower values would concentrate the probability mass function (PMF) for cases almost entirely on 0, while higher values effectively correspond to a Poisson distribution.

Applying the method to COVID-19 cases in US counties enabled investigation of changes in dispersion in relation to both observed trends and model expectations based on case counts (Fig 2). Periods of increased case count variability corresponded with decreases in *θ*, indicating that dispersion was dynamic. Changes in dispersion exhibited both expected and unexpected patterns of variation relative to standard theory. In some instances, *θ* varied inversely with incidence, consistent with standard epidemic theory, while in other periods, deviations from this expectation occurred, potentially signaling shifts in underlying transmission dynamics. Notably, significant changes in the dispersion parameter were observed during major epidemic transitions. For example, during the beginning of 2022, and at the end of the time series, when the pandemic was transitioning toward endemicity as the landscape of susceptibility was evolving [23]. The landscape of susceptibility was evolving as a larger proportion of cases involved reinfections. These findings underscore the complex behavior of the dispersion parameter, which not only varied with changes in case count regimes but also revealed departures from the model expectations described by Eq (1), which are consistent with changes in the underlying drivers of transmission.

**Fig 2.**
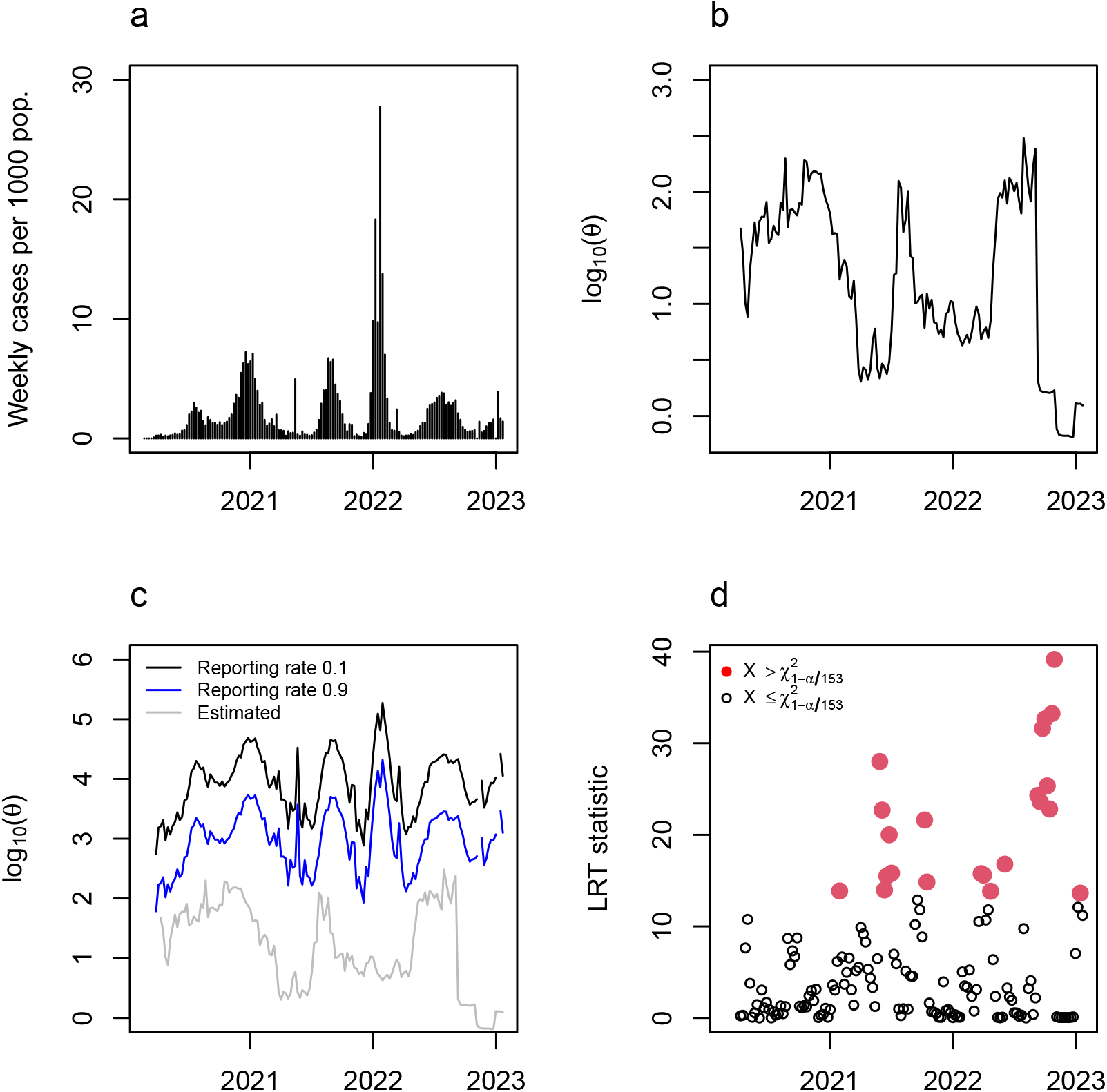
Dispersion analysis of weekly COVID-19 case data for Jefferson County, Alabama. Results for all counties are shown in Figure 3 a: Weekly reported COVID-19 incidence. b: Estimated dispersion parameter (*θ*) over time. c: Comparison of estimated dispersion (gray) with predicted values from the standard model *θ*_*t*+1_ = *C*_*t*_*/ρ*_*t*_, where *C*_*t*_ is reported cases and *ρ*_*t*_ is the reporting rate. Predictions are shown for fixed *ρ*_*t*_ = 0.1 (black) and *ρ*_*t*_ = 0.9 (blue), chosen to encompass the range of *θ* expected under variable *ρ*. d: Likelihood ratio test (LRT) statistic over time. Statistically significant changes in dispersion (red) correspond to p-values below the Bonferroni-corrected 5% threshold of a chi-square distribution with one degree of freedom.

Dispersion increased markedly around the peaks in incidence during the major 2022 wave, from late December 2021 to early February 2022 (Fig 3). This is in strong contrast to standard epidemic theory which predicts that dispersion should decrease as incidence rises. A high concentration of low *p*-values around peak incidence (Fig 3) corroborates widespread changes in *θ* across counties, reinforcing the statistical significance of this pattern. While these *p*-values should be corrected for multiple testing if used for inference rather than visualization, the overall trend suggests a systematic departure from theoretical expectations. Highly overdispersed patterns were also observed more frequently later in the time series, pointing to increasing heterogeneity in transmission, susceptibility, and reporting during the later phases of the pandemic. In both the 2022 wave and later in the pandemic, localized surges indicated by higher dispersion may have played a larger role in pandemic dynamics than expected, including potentially placing increased demand for surge capacity in hospitals and testing centers.

**Fig 3.**
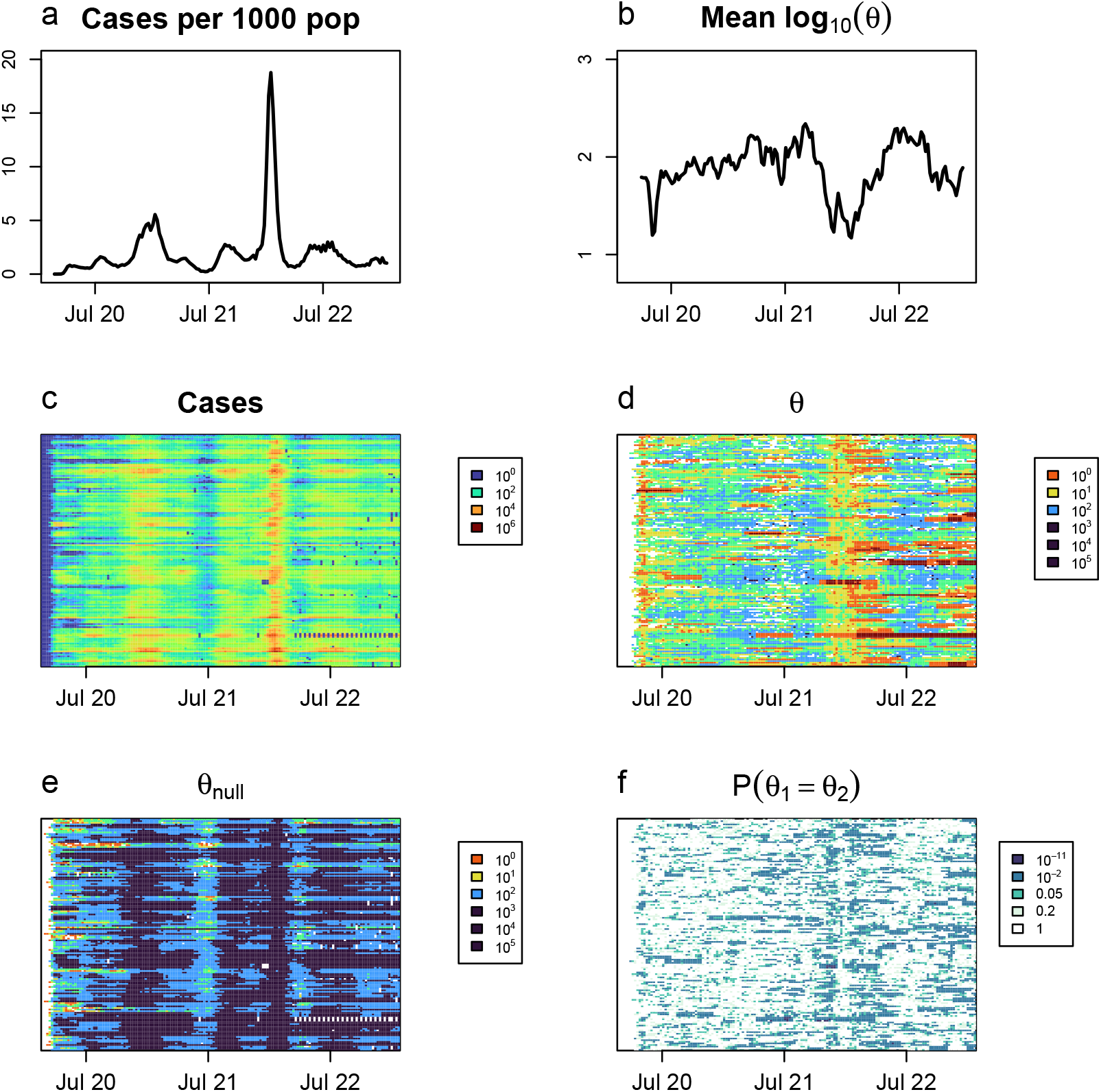
Incidence and dispersion between Jan 4, 2020 and March 18, 2023, in large counties in the US. a: Mean COVID-19 cases of the 144 US counties over time. b: Mean log_10_ *θ* of the 144 US counties over time. c: log_10_(*cases*) over time for each of the 144 counties. d: log_10_ *θ* over time for each of the 144 counties. e: Expected value of log_10_ *θ*_0_ under the null model, assuming a reporting rate of 0.5. f: LRT p-values over time for each location.

Our method forms part of a larger interest in investigating variability in infections as an important attribute of epidemic time series using novel metrics. For instance, burst-tree decomposition of time series has also facilitated computation of a burst-size distribution for a series given a specified time window [24], allowing comparison of variability within one location over time. Spatial variation in superspreading potential has been investigated through risk maps of superspreading environments [25], and future work could investigate the correspondence between dispersion in case count time series, as quantified here, and indicators of a high risk of superspreading, with the potential to further elucidate drivers of transmission risk across scales, and more finely resolve landscapes of susceptibility. Additionally, as population-wide disease control may be less effective than those which are focused to individuals in high-transmission contexts [1], identifying candidate time periods when transmission heterogeneity is high may catalyze the development of more effective control strategies, particularly those that connect vulnerable populations with resources at critical times. The finding that dispersion increased rather than decreased during the 2022 surge challenges theoretical expectations and suggests that fundamental assumptions about the scaling of transmission dynamics may require reevaluation. One hypothesis is that transmission heterogeneity could play a role in driving large surges, amplifying incidence beyond what homogeneous models predict. Future work could investigate whether bursts of highly clustered transmission events generate feedback that accelerates epidemic spread, which, if true, could refine predictive models of contagion dynamics in complex populations.

## Data Availability

The code used for the analysis reported in this paper can be accessed from
https://github.com/rachaelaber/dispersion. The data analyzed here are originally from
The New York Times and The United States Census Bureau, accessed via https://github.com/nytimes/covid-19-data/ and https://www2.census.gov/programs-surveys/popest/datasets/2020-2021/counties/totals/, respectively.

https://github.com/nytimes/covid-19-data/

https://www2.census.gov/programs-surveys/popest/datasets/2020-2021/counties/totals/

## Supporting Information

**S1 Text. Derivation of the relationship between the dispersion parameter and the mean crowding parameter**.

## Notes

### Competing Interest Statement

The authors have declared no competing interest.

### Funding Statement

RA was supported in part by the Achievement Rewards for College Scientists (ARCS) Foundation Oregon through an ARCS Scholar Program award and in part by the Association for Computing Machinery (ACM) SIGHPC Computational and Data Science Fellowship.
BDD was supported in part by a grant from the National Science Foundation under Award Number CBET-2200338, and by grants from the David and Lucile Packard Foundation.
The funders had no role in study design, data collection and analysis, decision to publish, or preparation of the manuscript.

### Author Declarations

The study used ONLY openly available human data that were originally located at https://github.com/nytimes/covid-19-data/ and https://www2.census.gov/programs-surveys/popest/datasets/2020-2021/counties/totals/.

