## Supplementary material for "Time-series modeling of epidemics in complex populations: detecting changes in incidence volatility over time": S1 Text

**S1 Text. Derivation of the relationship between the dispersion parameter and the mean crowding parameter.**

1. Let $m^*$ be mean crowding and $m$ be the mean.
$m^* = m + (\frac{\sigma^2}{m} - 1)$ (by definition of mean crowding)
$\implies m^* - m = \frac{\sigma^2}{m} - 1$

2. Let $\theta$ be the dispersion parameter and $\sigma^2$ be the variance of the negative binomial distribution.
$\sigma^2 = m + \frac{m^2}{\theta}$ (by definition of negative binomial variance)
$\implies \frac{\sigma^2}{m} = 1 + \frac{m}{\theta}$
$\implies \frac{m}{\theta} = \frac{\sigma^2}{m} - 1$

3. $\frac{m^*}{m} - 1 = \frac{1}{\theta}$ (by equating the left-hand sides of the results in 1 and 2 and dividing by $m$)
$\implies m^* = m + \frac{m}{\theta}$
